# Real World Effectiveness of Tixagevimab/cilgavimab (Evusheld) in the Omicron Era

**DOI:** 10.1101/2022.09.16.22280034

**Authors:** Benjamin Chen, Nina Haste, Nancy Binkin, Nancy Law, Lucy E. Horton, Nancy Yam, Victor Chen, Shira Abeles

## Abstract

**Background:** Pre-exposure prophylaxis for COVID-19 with tixagevimab/cilgavimab (T/C) received Emergency Use Authorization (EUA) based off of results from a clinical trial conducted prior to the Omicron variant. Its clinical effectiveness has not been well described in the Omicron era. We examined the incidence of symptomatic illness and hospitalizations among T/C recipients when Omicron accounted for virtually all cases.

**Methods:** We used the electronic medical record to identify patients who received T/C at our institution. Among these patients, we assessed for cases of symptomatic COVID-19 and associated hospitalizations before and after receiving T/C. We used chi square tests and Fishers exact p-values to examine differences between characteristics of those who got COVID before and after T/C prophylaxis.

**Results:** Of 1295 T/C recipients, 121 (9.3%) developed symptomatic COVID-19 before receiving T/C, and 102 (7.9%) developed symptomatic disease after receiving it. Among those with infection prior to T/C, 36/121 (29.8%) were hospitalized, including 8 (6.6%) admitted to the ICU. Among those with COVID-19 after receiving T/C, 6/102 (5.9%) were hospitalized but none required ICU admission. No COVID-related deaths occurred in either group. The majority of COVID-19 cases among those infected prior to T/C treatment occurred during Omicron BA.1 surge, while the majority of cases among post-T/C recipients occurred when BA.5 was predominant. Patients infected with COVID-19 prior to receiving T/C had received fewer vaccine doses and were less likely to receive COVID-19 therapeutics compared to those with COVID-19 after having received T/C.

**Conclusion:** We identified COVID-19 infections after T/C prophylaxis. Among persons eligible for T/C, COVID-19 illnesses occurring after T/C were less likely to require hospitalization compared to those with COVID-19 prior to T/C. In the presence of changing vaccine coverage, multiple therapies, and changing variants, the effectiveness of T/C in the Omicron era remains difficult to assess.

## Introduction

COVID-19 vaccines have dramatically reduced the severity of disease caused by SARS-CoV-2 infection.^1^ Immunocompromised patients, however, remain at risk for SARS-CoV-2 infections and, if infected, an increased risk of severe illness, hospitalization, and death. Although the overall mortality in immunocompromised patients with the SARS-CoV-2 Omicron variant has been shown to be lower than with prior variants, studies have shown that hospitalization and prolonged duration of symptoms have remained high for this vulnerable population.^2^

Tixagevimab/cilgavimab (AZD7442; Evusheld, subsequently referred to as T/C), which consists of AZD8895 and AZD1061 (AstraZeneca), two long-acting monoclonal antibodies, was shown in the PROVENT trial to reduce the risk of symptomatic COVID-19 infection by 83% compared with placebo.^3^ In early December 2021, the Food and Drug Administration (FDA) granted an Emergency Use Authorization (EUA) of T/C for pre-exposure prophylaxis for moderately to severely immunocompromised adults and adolescents, and for those not eligible for COVID-19 vaccination due to a history of a severe reaction.

The authorization of T/C coincided with the emergence of the Omicron variant, which resulted in previously effective monoclonal antibodies losing effectiveness.^4 5 6 7^ There is limited data available on the real-world effectiveness of T/C, especially in the Omicron era. T/C requires significant resources for administration due to its EUA status and associated institutional requirements for administering it. We therefore investigated COVID-19 infections and clinical outcomes in a cohort of immunocompromised patients who received T/C for pre-exposure prophylaxis within a single health system during a period when Omicron was the predominant circulating variant.

## Methods

The study was conducted at the University of California San Diego’s Health System (UC San Diego Health), a quaternary referral center serving a large population with many patients requiring complex subspecialty care. This system began administering T/C in January 2022 to patients according to the patient prioritization scheme set forth by the National Institute of Health’s COVID-19 Treatment Guidelines Panel.^8^ Patients initially received 300mg of drug, but the FDA recommended an increased dose for a total of 600mg on February 24, 2022, based on *in vitro* studies using the Omicron variant. ^9 10 11^ Patients who had received the 300mg dose were advised to receive an additional “catch-up” dose.

We reviewed the electronic medical records (EMR) of all patients who received T/C at our institution between January 1, 2022 and July 31, 2022. Records were matched with the health system’s COVID-19 testing data and with data on prescriptions for COVID-19 therapeutics including remdesivir (Veklury, Gilead), nirmatrelvir-ritonavir (Paxlovid, Pfizer), and neutralizing monoclonal antibodies (including REGEN-COV [casirivimab/imdevimab], sotrovimab, and bebtelovimab) from 10/01/2021 through 7/31/2022. Patients were considered to have had an episode of COVID-19 if they had a positive SARS-CoV-2 test recorded in their EMR or if they had been prescribed one of the therapeutic drugs between October 1, 2021, and July 31, 2022, through our health system. Cases in which therapeutic drug was prescribed were verified by chart review to ensure documentation confirming an active COVID-19 infection. We used EMR data to obtain each patient’s demographic and medical characteristics including history relevant to T/C eligibility, COVID immunization history, the clinical course of COVID-19 infection during the study period including whether infection resulted in hospitalization, intensive care unit stay, or death, dates and doses of T/C receipt, date of first positive COVID-19 test, and COVID-specific antiviral and monoclonal antibody therapeutics received.

We used these data to describe the characteristics of those who developed COVID-19 before receiving T/C or after receiving T/C. For the latter group, we calculated the interval between last dose of T/C and date of first positive test or prescription of COVID therapeutic. For those patients with multiple positive tests, we performed chart review to assess whether the repeat positive test was considered to have been consistent with a new infection. We used chi square tests and Fishers exact p-values to examine the difference between characteristics of those who got COVID before T/C and after T/C prophylaxis.

To evaluate the temporal distribution of COVID-19 infections in those who did and did not receive T/C prior to their illness, we plotted cases and hospitalizations by week of diagnosis and T/C status. To place the cases in the context of the COVID-19 patterns in the community and which variants were circulating in the San Diego area during the same time interval, we used data from the SEARCH GitHub repository (San Diego Wastewater Surveillance - SEARCH (searchcovid.info)) to develop a graph of mean weekly wastewater concentrations of SARS-CoV-2 by variant type.

To examine the impact of T/C on rates of hospitalization, we calculated the proportion of cases who required hospitalization among patients who received T/C before developing COVID-19 and those who developed it after receiving T/C. We calculated rate ratios, 95% confidence intervals and two-tail Fischer exact p-value. For this analysis, we also examined the rates by underlying condition (hematologic malignancy including bone marrow transplantation, solid organ transplantation, and other, which included patients with underlying immunosuppression such as rheumatologic conditions or advanced AIDS).

This study was determined by the UC San Diego Institutional Review Board to be exempt from Institutional Review Board requirements under categories 45 CFR 46.104(d), category 4.

## Results

### General patient characteristics

From January 1, 2022, through July 31, 2022, UC San Diego Health administered pre-exposure T/C prophylaxis to 1295 unique patients, of whom approximately 9.2% received it in January (the majority of which received “catch-up” doses in March), 18.8% in February, 17.1% in March, 23.3% in April, 14.0% in May, and the remaining 17.6% between June and July 2022.

The median age of the patients who received T/C was 59 years (range 18-99 years) and 57.5% were male, 42.4% were female, and 0.1% were non-binary. A total of 37.2% were solid organ transplant recipients, 47.7% had received bone marrow transplants or had hematologic malignancies, and 15.1% had other qualifying conditions (including on active chemotherapy, advanced HIV/AIDS, on significant immunosuppression for an autoimmune disorder, etc).

### Characteristics of patients who developed COVID-19

Of the 1295 T/C recipients, 223 met the COVID-19 case definition, of whom 121/223 (54.3%) became ill with COVID-19 before receiving T/C and 102/223 (45.7%) developed breakthrough COVID-19 infection after receiving pre-exposure prophylaxis (Table). A total of 168/223 (75.3%) were identified based on test results, and the remaining 55/223 (24.7%) on the basis of having received COVID-19 therapy. The median elapsed time to COVID-19 positivity was 80 days following T/C administration (range 1 to 204 days).

**Table:** Characteristics of recipients of tixagevimab-cilgavimab (T/C) who developed COVID-19 infection between October, 1 2021 and July 31, 2022

| Patient Characteristic | Patients with COVID-19 infection prior to receipt of T/C (n = 121) | Patients with breakthrough COVID-19 infection following receipt of T/C (n = 102) | p-value |
| --- | --- | --- | --- |
| **Median age [range], years** | 54.5 [18 – 79] | 60.5 [25 – 99] | $p = 0.01$ |
| **Patient gender** |  |  |  |
| Male (%) | 66 (54.5) | 58 (56.9) |  |
| Female (%) | 55 (5.5) | 44 (43.1) |  |
| **Underlying conditions** |  |  |  |
| Solid Organ Transplantation (%) | 49 (40.5) | 35 (34.3) |  |
| Hematologic Malignancy (%) | 37 (30.6) | 28 (27.5) |  |
| Bone Marrow Transplantation (%) | 16 (13.2) | 28 (27.5) |  |
| Autoimmune (%) | 9 (7.4) | 3 (2.9) |  |
| Neurologic (%) | 4 (3.3) | 3 (2.9) |  |
| Solid Tumor Malignancy (%) | 3 (2.5) | 2 (2.0) |  |
| HIV-AIDS (%) | 3 (2.5) | 2 (2.0) |  |
| Other (%) | 0 (0.0) | 1 (1.0) |  |
| **Number (%) of vaccine doses received prior to COVID diagnosis<sup>†</sup>** |  |  |  |
| 0 | 19 (15.7) | 3 (3.0) | $p < 0.001$ |
| 1 – 2 | 33 (27.3) | 25 (24.8) |  |
| 3 | 61 (50.4) | 36 (35.6) |  |
| 4 | 8 (6.6) | 37 (36.6) |  |
| **Therapeutics received (%)** |  |  |  |
| Nirmatrelvir-Ritonavir | 6 (5.0) | 40 (39.2) | $p = 0.01$ |
| Bebtelovimab | 0 (0.0) | 34 (33.3) |  |
| Remdesivir | 22 (18.2) | 17 (16.7) |  |
| Sotrovimab | 56 (46.3) | 2 (2.0) |  |
| Casirivimab-Imdevimab | 10 (8.2) | 0 (0.0) |  |
| None | 27 (22.3) | 9 (8.8) |  |
| **Number of patients hospitalized (%)** | 36 (29.8) | 6 (5.9) | $p < 0.001$ |
| **Number of patients with ICU stay (%)** | 8 (6.6) | 0 (0.0) | $p = 0.01$ |
| **Hospital length of stay, days<sup>‡</sup> (%)** |  |  |  |
| 1 – 7 | 17/36 (47.2) | 3/5 (60.0) |  |
| 8 – 14 | 7/36 (19.4) | 1/5 (20.0) |  |
| 15 – 30 | 3/36 (8.3) | 1/5 (20.0) |  |
| 31 – 60 | 4/36 (11.1) | 0 (0.0) |  |
| ≥ 61 | 5/36 (13.9) | 0 (0.0) |  |
| **Number of deaths from COVID-19 (%)** | 0 (0.0) | 0 (0.0) |  |

†One patient's vaccine history was not documented within the electronic medical record ‡One patient was hospitalized at an outside institution and medical records were unavailable to us
**Abbreviations:**
ICU = Intensive Care Unit
HIV-AIDS = Human Immunodeficiency Virus – Acquired Immunodeficiency

The distribution of cases over time in our study cohort is shown in Figure 1A. Few cases were identified between October and mid-December, after which there was a sharp increase in cases coinciding with the rapid emergence of the Omicron variant (Figure 1C). The number of cases peaked in the first week of January, falling to a more baseline level by the end of the month. All of the cases before and during the winter peak were among patients who had not yet received T/C. Figure 1B demonstrates a parallel rise in hospitalization during the winter peak, with all cases occurring among those who had not yet received T/C. The local circulating strain was initially Omicron BA.1, which was rapidly replaced by BA.1.1 (Figure 1C). In late April 2022, cases began to rise again, with wastewater data showing increasing levels of Omicron BA2.12 and subsequently BA.5.

**Figure 1A.**
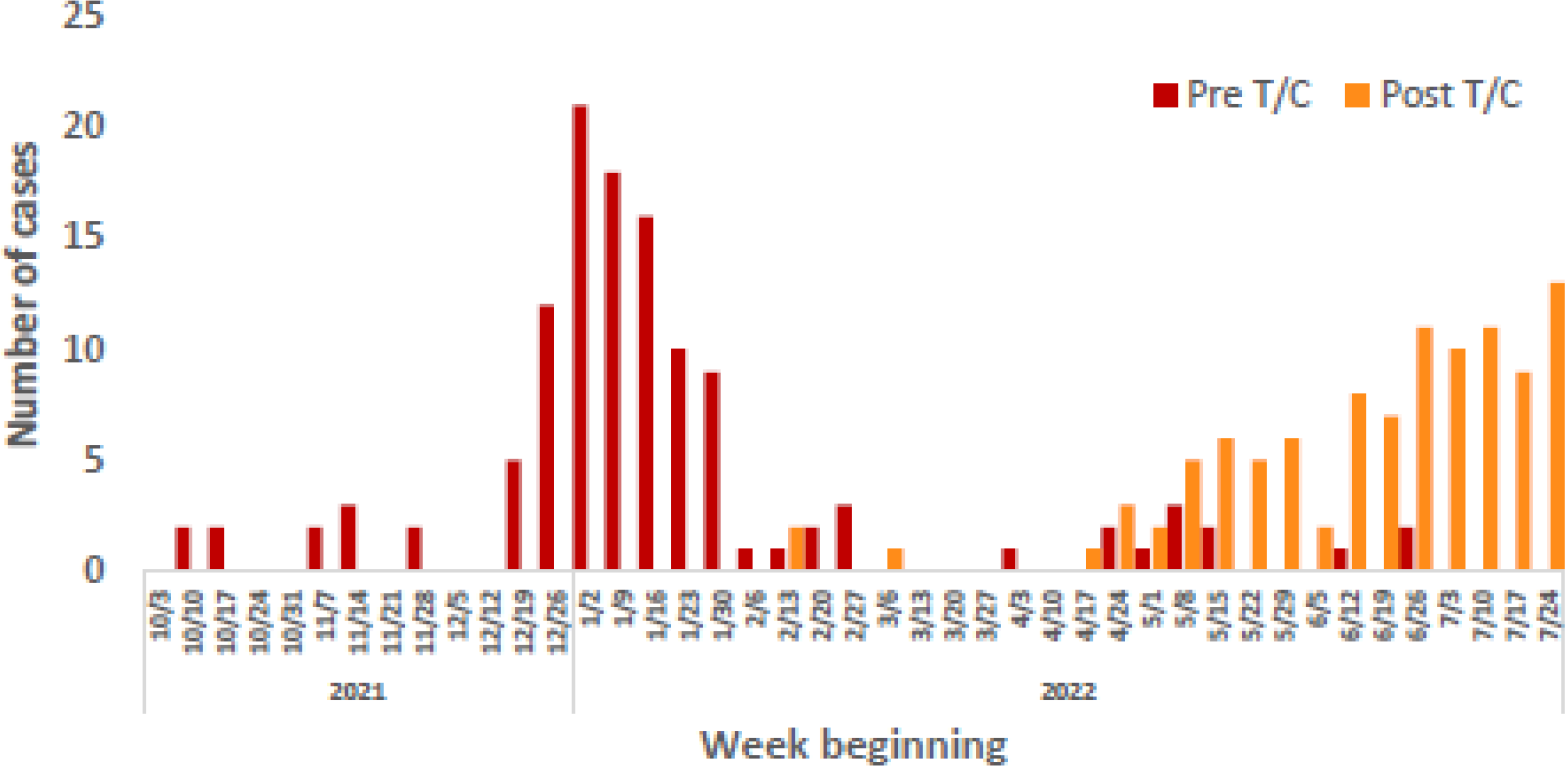
Cases of COVID-19 by week and T/C status at the time of COVID-19 diagnosis, UC San Diego Health, October 2021 - July 2022

**Figure 1B.**
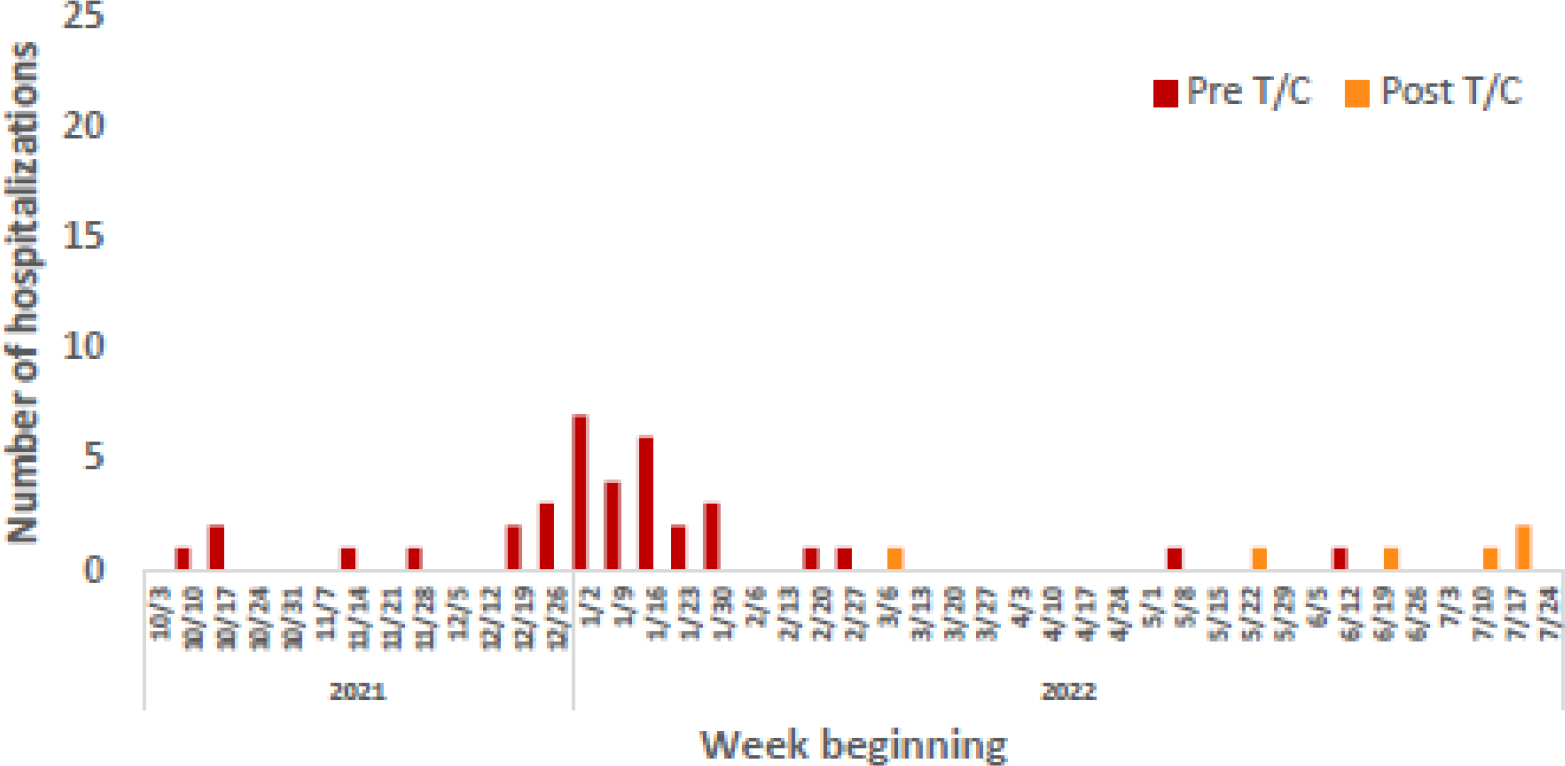
Hospitalizations for COVID-19 by week and T/C status at the time of COVID-19 diagnosis, UC San Diego Health, October 2021 - July 2022.

**Figure 1C.**
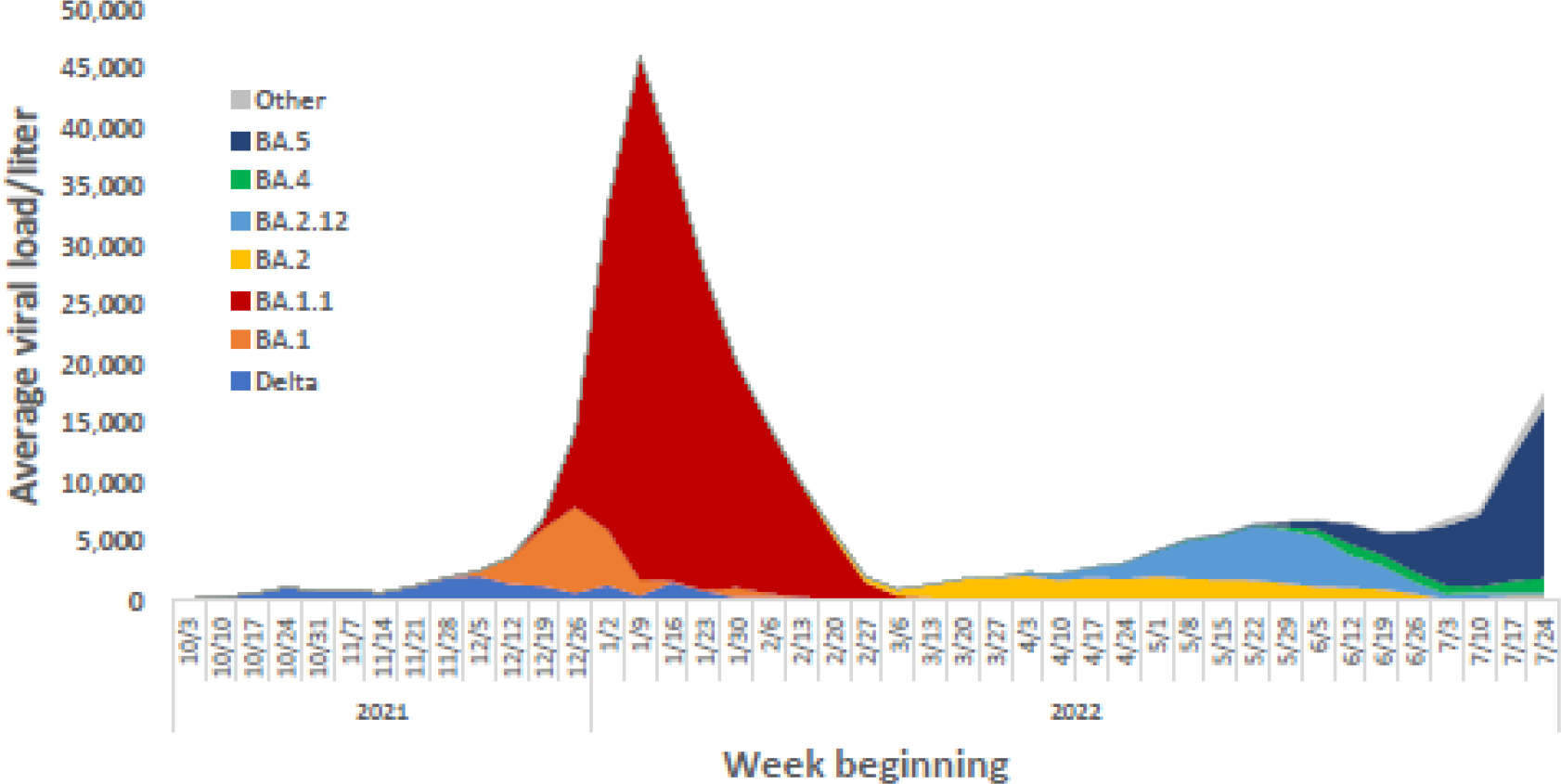
Average weekly SARS CoV-2 wastewater viral loads/liter by variant, San Diego County, October 2021 – July 2022.

We compared the 121 patients who developed COVID-19 prior to receiving T/C (pre-T/C) with the 102 patients who developed it after receiving T/C (post T/C; Table). The pre-T/C patients infected with COVID-19 were significantly younger than the post-T/C patients infected with COVID-19 (median 54.5 years versus 60.5 years; Mann-Whitney Wilcoxson two-sample test p = 0.01). Similar proportions of the pre-T/C and post-T/C COVID-19 cases were male (54.6% versus 56.9%). Among the pre-T/C cases, 53/121 (43.8%) had hematologic malignancies or were bone marrow transplant recipients, 49/121 (40.5%) were solid organ transplant recipients, and the remaining 19/121 (15.7%) had other forms of immunosuppression. Among the post-T/C patients, 56/102 (54.9%) had hematologic malignancies or were bone marrow transplant recipients, 35/102 (34.3%) were solid organ transplant recipients, and 11/102 (10.8%) had other forms of immunosuppression. The differences in distribution by gender and underlying condition were not statistically significant.

Vaccine status and treatment with therapeutic agents also differed between pre-T/C and post T/C COVID-19 cases. A higher proportion of the pre-T/C cases had not received any doses of COVID-19 vaccine (15.7%, versus 3.0% for the post-T/C cases); conversely the post-T/C cases were more likely to have received three or four doses (72.2% versus 57.0%). The difference in number of doses was statistically significant (p < 0.001). The majority (94/121; 77.7%) of the pre-T/C cases were treated with a therapeutic agent, most commonly sotrovimab (56/121; 46.3%) and remdesivir (22/121; 18.2%), and 6/121 (5.0%) received nirmatrelvir-ritonavir, although the percentage of the post-T/C receiving a therapeutic agent was significantly higher (91.2%; p = 0.01) than the pre-T/C. In the post-T/C group, the most common therapeutic agents used included nirmatrelvir-ritonavir (40/102; 39.2%), bebtelovimab (34/102; 33.3%), and remdesivir (17/102; 16.7%).

### Hospitalization

Thirty-six of the 121 (29.8%) patients who developed COVID-19 prior to receiving T/C were hospitalized, with a median hospital length of stay of 8 days (range 1 to 119 days). None of the patients died, though 8/121 (6.6%) required an intensive care unit stay. Two patients underwent lung transplantation as a result of severe acute respiratory distress syndrome due to COVID-19 and subsequently became eligible for, and received, T/C after transplantation.

Six of 102 patients were hospitalized with COVID-19 after having received T/C (5.9%) with a median hospital length of stay of 7 days (range 3 to 16 days). Four of the six patients were solid organ transplant recipients, one patient had hematologic malignancy, and one patient was on immunosuppressive therapy for an underlying autoimmune disease. All of the hospitalized patients presented with symptoms consistent with a viral respiratory infection including fever, cough, weakness, and myalgia. Three of the hospitalized patients exhibited hypoxemia, and one of the patients also had a *Legionella* co-infection. No patients in this population required an intensive care unit stay or died.

The rate of hospitalization among those receiving T/C pre-exposure prophylaxis was one-fifth the hospitalization rate of those who developed COVID-19 prior to receiving T/C (rate ratio (RR) = 0.20; 95% confidence intervals (CI) = 0.09-0.45; p <0.001). The hospitalization rate for post-T/C patients with a hematologic malignancy or bone marrow transplant who developed COVID-19 was less than one-tenth that of those who had not yet received T/C (RR = 0.09; 95% CI = 0.01-0.64; p = 0.002). For patients who had received solid organ transplants, the corresponding values were RR = 0.27; 95% CI = 0.10-0.71; p = 0.003; for patients with all other conditions, the RR was 0.43, with 95% CI = 0.05-3.4; p = not significant.

## Discussion

We identified many COVID-19 cases among high-risk patients in our health system who had received T/C for pre-exposure prophylaxis. However, when we compared those who had contracted COVID-19 prior to receiving T/C prophylaxis with those infected after, patients with infection after receiving T/C were significantly less likely to be hospitalized for COVID-19.

Our findings differ from those of the initial clinical trial of T/C which had demonstrated an 83% reduced risk of symptomatic infection. However, that trial was conducted prior to the emergence of Omicron, which has been the predominant circulating variant worldwide since December 2021. Indeed, other monoclonal antibodies have been shown to be ineffective against the Omicron variants. ^9 10 11^ *In vitro* studies of cilgavimab, one of the two components of T/C, suggested it would maintain some efficacy against the emerging Omicron variants, but at a higher concentration than was used in the PROVENT trial. In February 2022 the FDA recommended dose of T/C should be doubled based on these predictions.

It is difficult to assess the impact of T/C prophylaxis on the low hospitalization rate in our cohort. Almost all who had COVID-19 after receiving prophylactic T/C were also offered additional treatment for symptomatic COVID-19, including updated monoclonal antibodies or antivirals, and nearly three-quarters had received at least three COVID-19 vaccine doses, including 37% who had gotten four. In addition to the availability of additional vaccine doses and a greater range of effective COVID-19 therapeutics over time, there have also been rapidly changing subvariants of varying severity and increasing immune evasiveness, further complicating the interpretation of therapeutic effectiveness. Monoclonal antibodies targeting the regions of the spike protein of the SARS CoV-2 virus under evolutionary pressure have not been able to be studied at a pace that meets the speed of the evolution of variants in the COVID-19 pandemic.

Our study has several limitations. First, although we used multiple data sources to identify cases, patients receiving COVID-19 care at other health care systems may not have been captured in our data. Second, because the limitations imposed by the use of administrative data, it was not possible to conduct a more rigorous analyses that might have allowed us to determine the relative contributions of T/C, underlying patient conditions, vaccine, circulating variants, and antiviral and monoclonal antibody treatment to the risk of developing COVID-19. Finally, we did not have complete variant data at the individual patient level to better assess risk of hospitalization by variant.

Despite these limitations, we believe our findings demonstrate that high-risk patients who have received T/C are still developing symptomatic COVID-19 infections in the Omicron era. Our results raise questions about the ongoing value of T/C pre-exposure prophylaxis. This long-acting antibody combination requires significant resources for administration to highly vulnerable patients. Given the many rapidly changing aspects of the COVID-19 pandemic, including the evolution of variants and subvariants, the protection offered by additional vaccine doses, and the development of effective antivirals and therapeutic monoclonal antibodies, we are unable to truly assess the contribution of T/C to the improved outcomes among patients over time without a rigorous clinic trial. Such a trial, however, would be difficult to perform and would run the risk of being rapidly outdated. As a result, decisions regarding the significant resources required to administer this drug unfortunately continue to depend on data from *in vitro* studies and expert recommendations.

## Data Availability

Data cannot be shared publicly because of patient privacy regulations. De-identified data could be made available with institutional oversight to ensure data sharing is consistent with strict institutional deidentification guidance.

## Acknowledgements

We thank Shobha Kolan, Thomas Hatch, and Smruthi Karthikeyan for their contributions to this study. We acknowledge the work performed by the UC San Diego Health clinical team who provided tixagevimab/cilgavimab to patients. No-one received compensation for their roles in this study.

## Author Contributions

BC contributed to data curation, investigation, formal analysis, writing and editing, NH contributed to data curation, formal analysis, writing, and editing, NB contributed to investigation, formal analysis, writing, and editing, NL contributed to methodology, project administration, writing, and editing, LH contributed to methodology, project administration, writing, and editing, NY contributed to data curation, methodology, project administration and editing, VC contributed to investigation, project administration, and editing, and SRA contributed to conceptualization, project administration, methodology, project administration, writing, and editing.

